# Pudendal nerve stimulation recruits the urethra during awake human cystometry

**DOI:** 10.64898/2026.02.18.26346494

**Authors:** Amador C Lagunas, Po-Ju Chen, Tim M Bruns, Priyanka Gupta

## Abstract

**Objective:** This study aimed to characterize the activation of lower urinary tract (LUT) targets in response to pudendal nerve stimulation (PNS) in awake human participants.

**Materials and Methods:** In this single center study, recruited participants had an implanted pudendal neurostimulator for treatment of their symptoms including overactive bladder, incontinence, urinary retention, and/or pelvic pain. Participants came in for a modified urodynamic study where a multichannel manometry catheter was placed in the lower urinary tract alongside a dual sensor urodynamics catheter. The bladder was filled and after each participant expressed a strong desire to void, PNS was applied and LUT pressures were measured. Participants attempted voids with the catheters in place to characterize LUT behavior and voiding efficiency with and without stimulation.

**Results:** The study consisted of 15 sessions with 2 male participants and 11 female participants, with 2 female participants completing two sessions. PNS-driven urethra pressure changes were observed in 50% (31/62) of high frequency (30-33 Hz) trials and 34% (24/71) of low frequency (2-3.1 Hz) trials. There was a significantly higher maximum tolerable stimulation amplitude for low frequency stimulation when compared to high frequency stimulation (p = 0.015). During high frequency stimulation, the maximum observed pressure change occurred significantly more often at the proximal urethra than the distal urethra (p = 0.007). In one participant there were four instances of stimulation driven bladder contractions with an average pressure change of 24.3 cmH_2_O (standard deviation = 10.5). There was not a significant difference in voiding efficiency or maximum flow rate with and without stimulation (p = 0.76 and p = 0.45, respectively).

**Conclusions:** PNS can affect LUT pressures at tolerable stimulation amplitudes. The absence of an effect of PNS on voiding characteristics suggests a similar mechanism of action as sacral neuromodulation.

## Introduction

Pudendal nerve stimulation (PNS) is an off-label clinical treatment for incontinence, retention, and chronic bladder pain that has been in use for 20 years^1–3^. However, there is still little information on how pudendal stimulation directly affects the lower urinary tract in human subjects^4,5^. While the relationship between PNS and lower urinary tract activation has been well studied in animal models^6–8^, most findings have not been verified in human subjects. An improved understanding of the effects of PNS in humans could lead to improvements for a variety of lower urinary tract dysfunctions.

The pudendal nerve is a mixed nerve that innervates the perineum, genitals, and external urethra sphincter^9^. While PNS can cause external urethra contractions in preclinical experiments^6^ and in humans under anesthesia^10^, it’s not clear if PNS can generate urethra contractions without bothersome pain or discomfort in awake humans. If feasible, on demand urethra contractions in humans could help treat stress urinary incontinence stemming from inadequate urethra pressure.

As a mixed nerve, the pudendal nerve contains sensory afferents^9^ that contribute to bladder function through spinal reflex circuits^11,12^. Animal studies have shown that PNS can lead to bladder contractions^13,14^ and there are reported cases of PNS evoked bladder contractions occurring in individuals with spinal cord injury^15,16^. However, there have not been any reported instances of PNS evoked bladder contractions in non-spinal cord injured individuals. The ability to generate bladder contractions with PNS could help people with detrusor underactivity manage their symptoms and regain some voiding function.

PNS can improve symptoms for patients with overactive bladder and retention, but the mechanism that causes symptom improvement is unclear. The effectiveness of PNS is counter intuitive as overactive bladder and retention are issues with contradictory bladder symptoms.

Preclinical studies have shown that PNS can directly affect voiding and bladder function^7,17^ but no studies have looked at the direct effects of PNS on voiding characteristics in human participants. A study on voiding performance with and without stimulation may improve our understanding of how PNS improves patient symptoms and help improve future therapies.

In this work we investigated direct and indirect effects of PNS on the lower urinary tract (LUT) in participants with an implanted pudendal neurostimulator. We examined the effect of PNS on urethra pressures at multiple locations, attempted to elicit reflex bladder contractions, and compared LUT dynamics during voluntary voiding with and without PNS.

## Materials & Methods

Recruited participants had an implanted pudendal neurostimulator for treatment of symptoms including overactive bladder, pelvic pain, incontinence, and/or urinary retention.

A visualization of the experimental setup is shown in Figure 1. Each participant was seated, in a reclined position, in a standard urodynamics suite. We recorded pressures in the bladder and urethra with a 2.75 mm diameter manometry catheter (MSC-3886: Manoscan, Medtronic, MN, USA) with solid-state pressure sensors at a 7.5 mm center to center distance between adjacent sensors. We estimated the position of each pressure sensor in the LUT from the pressures recorded by the manometry catheter and from the number of the sensor located outside of the body. We identified the proximal urethra sensor as the sensor with the highest pressure outside of the bladder. For female participants we set the distal urethra sensor as the first sensor inside the body and for male participants we set the distal urethra sensor as three sensors away distally from the proximal urethra sensor. We obtained bladder pressures from the manometry sensor that was closest to the tip of the catheter. We also followed standard cystometry practice with a 7 French air-charged dual sensor catheter (CAT880: Laborie, NH, USA) placed in the LUT to infuse saline into the bladder and to record secondary bladder and urethra pressures, which were not used here. An abdominal catheter (CAT875: Laborie, NH, USA) was placed in the rectum (if possible) or else in the vagina to record abdominal pressure. A pair of patch electrodes were placed on opposite sides of the anal sphincter to record anal sphincter activation and electrical stimulation artifact. A uroflowmeter was placed underneath each participant to measure urine flow.

**Figure 1:**
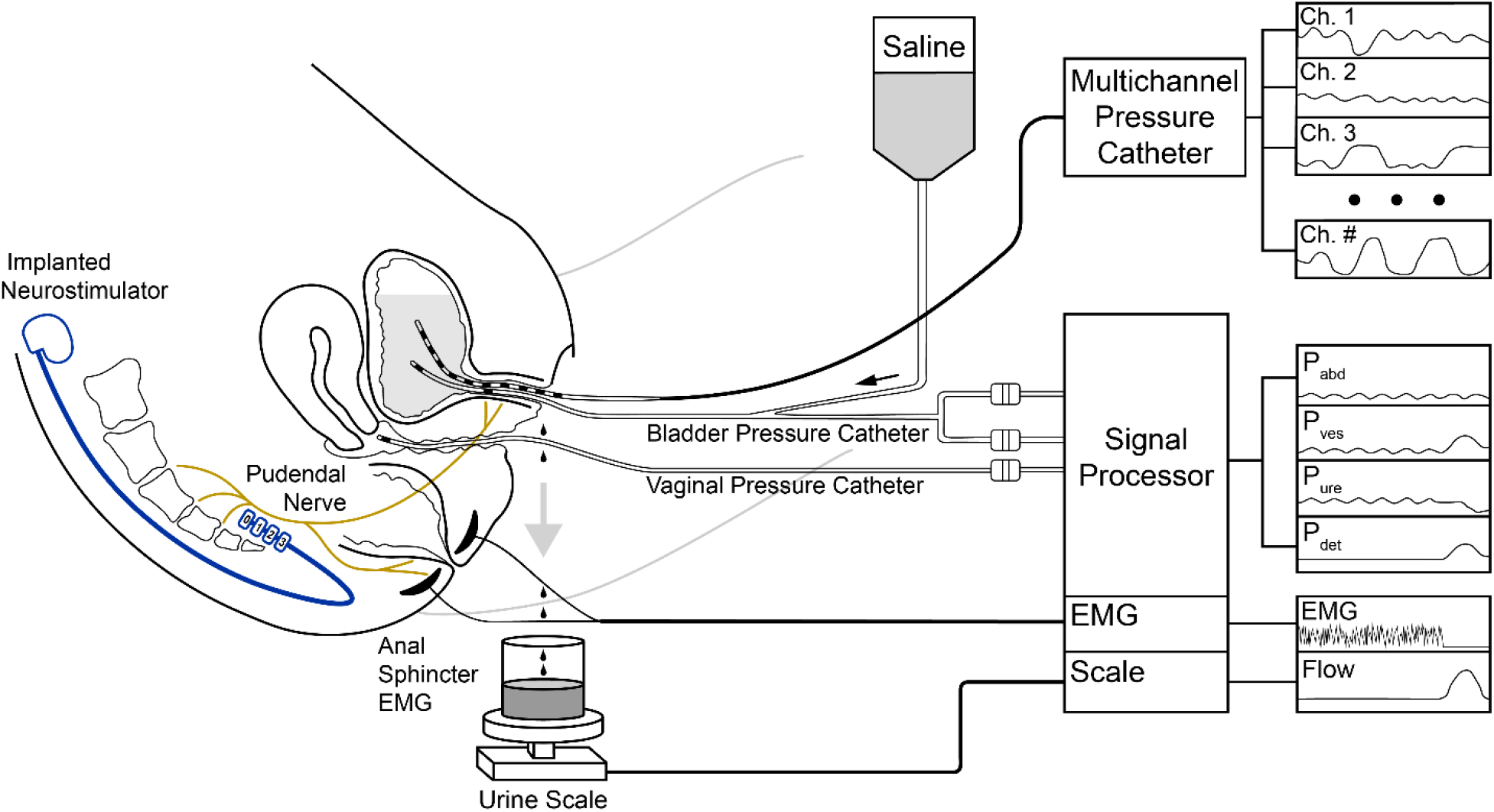
Experimental setup diagram. With participants in a reclined position, a multi-channel catheter was inserted into the urethra to measure urethra and bladder pressures. The multichannel catheter was placed alongside a urodynamics catheter which was used to infuse saline into the bladder. A urodynamics system was used to record bladder, urethra, and abdominal pressures, anal sphincter EMG, and the voided volume. This figure was modified from Karram and Blaivas^32^.

In each testing session, we filled the bladder with saline while PNS was turned off until the participant reported a strong desire to urinate. We then performed stimulation trials during which we evaluated a different monopolar or bipolar electrode configuration (e.g., electrode 0-(cathode) and electrode 3+ (anode); see Figure 1 for electrode labels) with 2-3.1 Hz, referred to as low frequency here after, and 30-33 Hz, referred to as high frequency here after. In preclinical studies low frequency stimulation (2-5 Hz) and high frequency stimulation (20-40 Hz) can cause bladder inhibition and excitation^18–21^ so these stimulation frequency ranges were chosen to see if similar results could be observed in humans. For each trial, the amplitude was increased to the maximum tolerable stimulation amplitude (MTSA) for that configuration and frequency and kept there for at least ten seconds. After the stimulation trials, participants attempted to void and we applied PNS with their home program or a program that recruited the pelvic floor at the stimulation amplitude they used at home. After the first void, if the participant agreed, we conducted another filling cycle with additional stimulation trials and then a second void attempt without PNS.

For low frequency stimulation trials, the urethra pressure contraction amplitude was calculated by averaging the peak height for five stimulation-driven contractions. For high frequency stimulation trials, the urethra pressure contraction amplitude was calculated by taking the difference between the maximum pressure during stimulation and a five second average baseline before stimulation was applied. When there were multiple sensors in the distal or proximal urethra, the sensor with maximum pressure change was used in the analysis. The bladder pressure change during voiding was calculated as the difference between the mean bladder pressure in the ten-second interval surrounding the peak flow rate and in the ten-second interval before the void was initiated. Voiding efficiency was expressed as the ratio of voided volume to infused volume. If more fluid was voided than infused then efficiency was set at 100%^22^.

We used the Shapiro-Wilk test to check if data was normally distributed. If the data was normally distributed, then it is reported as mean with standard deviation (SD) otherwise data is reported as median with interquartile range (IQR). After confirming normality of the data, we used a Welch two sample t-test to check for a significant difference between groups. A paired t-test was used to compare bladder pressure changes during voids. A Wilcoxon rank sum test was used if data was non-normal. To evaluate statistical significance, α was set to 0.05.

## Results

We collected data from 13 participants over 15 cystometry sessions. One participant was reenrolled after receiving a new pudendal stimulator on the opposite nerve. Another participant was reenrolled after a fall that affected symptom relief from the implanted stimulator. Individual participant demographic data and diagnoses are shown in Table 1.

**Table 1.** Participant demographic information.

| Participant | Sex | Age (years)<br>(5-year intervals) | Body Mass Index<br>(kg/m<sup>2</sup>) | Condition(s) |
| --- | --- | --- | --- | --- |
| 1002<sup>‡</sup> | F | 61-65 | 22.3 | OAB, PP |
| 1008 | F | 56-60 | 25.8 | Ret, PP |
| 1010 | F | 76-80 | 30.4 | OAB, Ret, PP |
| 1012 | F | 51-55 | 21.3 | OAB, Ret |
| 1016 | F | 46-50 | 30.7 | OAB, PP |
| 1017* | F | 61-65 | 28.0 | OAB, PP |
| 1019 | F | 56-60 | 31.6 | OAB |
| 1021* | F | 61-65 | 26.4 | OAB, PP |
| 1022 | F | 56-60 | 36.7 | OAB, PP |
| 1024 | F | 51-55 | 26.2 | OAB, PP |
| 2001 | M | 31-35 | 35.9 | OAB, PP |
| 2003 | F | 66-70 | 31.2 | PP |
| 2004 | M | 51-55 | 30.1 | Ret |
| 2005 | F | 66-70 | 31.8 | PP |
| 2006<sup>‡</sup> | F | 61-65 | 23.0 | OAB, PP |
| N = 15 | F=13 | 59 ± 11 | 28.8 ± 4.6 |  |

\* Participant 1017 was reenrolled as 1021 after receiving a new implant. <sup>‡</sup> Participant 1002 had a second study visit and was (re)enrolled as 2006. Summary statistics are shown as mean ± standard deviation. OAB: Overactive Bladder; Ret: Urinary Retention; PP: Pelvic Pain.

Across all low and high frequency trials at full bladder volumes, for 11 cystometry sessions with voltage-controlled stimulators, the MTSA was not normally distributed (p = 7.4e-9). Trials at low frequency had a median MTSA of 2.4 V (IQR = 2.0) and high frequency trials had a median MTSA of 1.9 V (IQR = 1.7). There was a significant difference (p = 0.015) between the median MTSA for low and high frequency stimulation. Trials at low frequency with observed contractions had a median MTSA of 1.9 V (IQR = 0.6) which was not significantly different (p = 0.10) than trials without observed contractions which had a median MTSA of 3.5V (IQR = 3.3). Trials at high frequency with observed contractions had a median MTSA of 1.9V (IQR = 0.6) which was not significantly different (p = 0.69) than trials without observed contractions which had a median MTSA of 1.9 V (IQR = 2.7).

We observed urethra contractions in response to stimulation in 11 sessions. Across 62 low frequency trials in nine sessions, 31 trials had observable contractions at the MTSA. The number of responsive trials and the median distal and proximal pressure change for each participant are shown in supplemental Table 1. When urethra contractions were observed, the maximum pressure change occurred at the distal urethra 19 times and at the proximal urethra 12 times, with no significant difference (p = 0.28) in the proportion of maximum pressure at each location. For low frequency stimulation, the median stimulation evoked contraction amplitude was 2.3 cmH2O (IQR = 3.1) at the distal urethra and 2.6 cmH2O (IQR = 5.9) at the proximal urethra with no significant difference (p = 0.53) between locations. Figure 2A illustrates the relationship between the location in the urethra and the observed contraction amplitude during low frequency trials. For high frequency stimulation there were urethra contractions in 24 out of 71 trials in seven sessions. The maximum pressure change occurred in the distal urethra 5 times and the proximal urethra 19 times, with a significant difference (p = 0.007) in the proportion of trials with a larger contraction in the proximal urethra. For high frequency stimulation the median stimulation evoked contraction amplitude was 17.5 cmH2O (IQR = 29.9) at the distal urethra and 33.8 cmH2O (IQR = 41.3) at the proximal urethra with a significant difference (p=0.013) between locations. Figure 2B illustrates how pressure changes with urethra location for high frequency trials.

**Figure 2:**
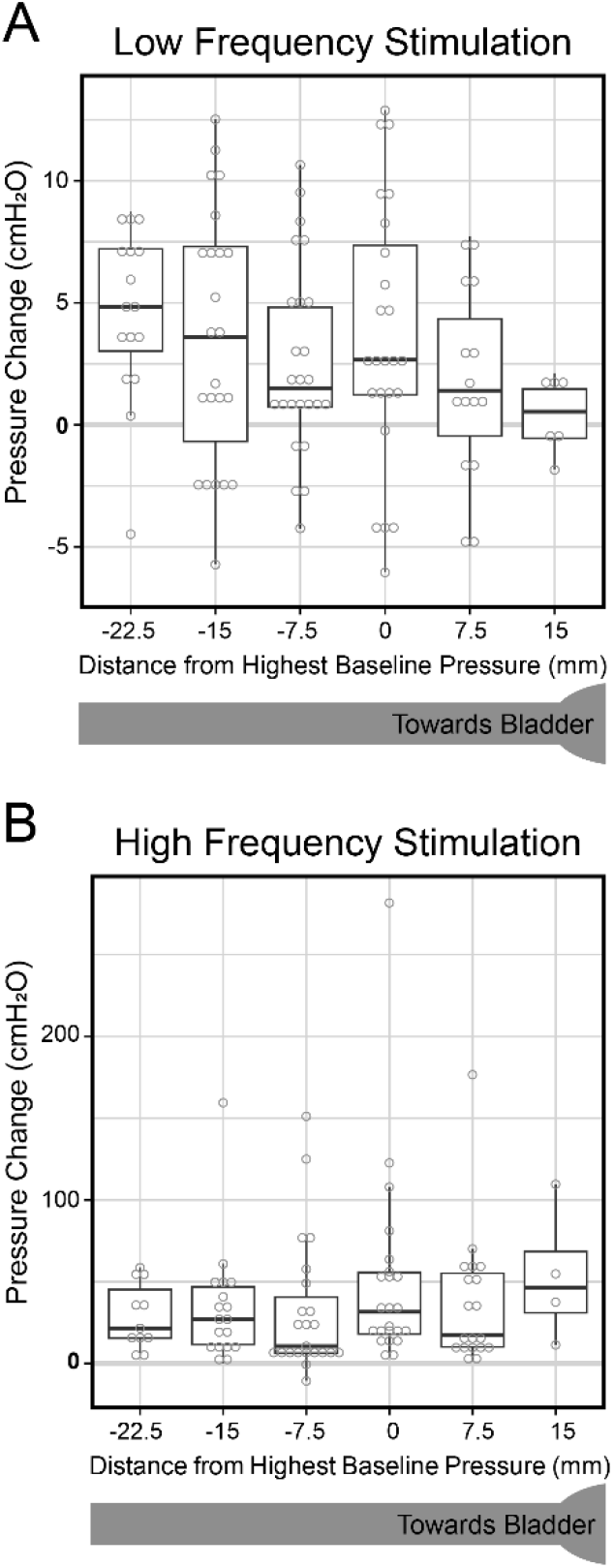
Stimulation driven urethra pressure changes at maximum tolerable stimulation amplitude across participants. The urethra position is labeled according to each sensors relative position to the sensor with the greatest baseline pressure. A) Lower urinary tract pressure changes during stimulation at low frequency. Pressure change is calculated per stimulation pulse. The Y-axis is labelled in increments of 5 cmH20. B) Lower urinary tract pressure changes during stimulation at high frequency. Pressure change is the maximum difference from the baseline pressure during a tetanic contraction. The Y-axis is labelled in increments of 100 cmH20.

We observed four instances of stimulation driven bladder contractions in one participant (2001). The pressure traces for two example bladder contractions are shown in Figure 3. The mean magnitude of the bladder contraction was 24.3 cmH2O (SD = 10.5). The bladder contractions were observed with 2.1 Hz and 31 Hz stimulation, with two different contacts, and at pulse widths of 210 and 450 µs. All contractions occurred at bladder volumes above 70 percent with monopolar stimulation or bipolar stimulation and with a MTSA at or below 2 V.

**Figure 3:**
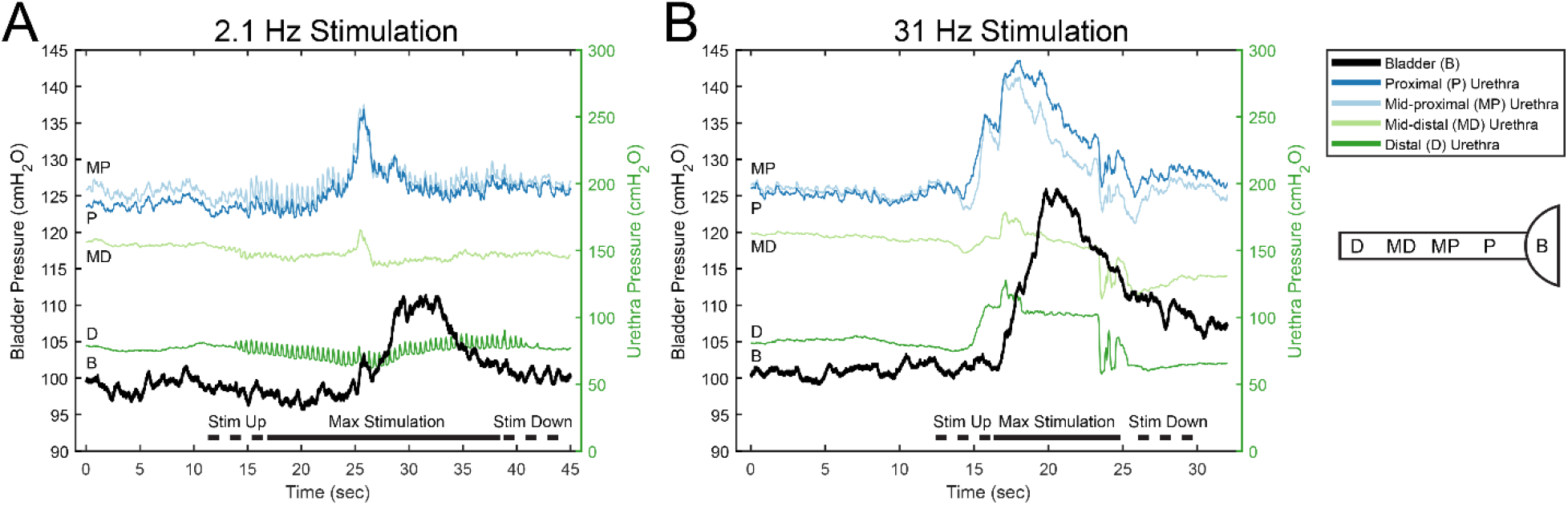
LUT pressures from example trials with bladder contractions in participant 2001. A) A trial with stimulation applied at 2.1 Hz and a maximum tolerable stimulation amplitude of 2V. B) A trial with stimulation applied at 31 Hz and a maximum tolerable stimulation amplitude of 1.3V. Note the different timing dynamics of bladder pressure rise or fall as compared to urethra pressure changes.

There were five participants who voided both with and without stimulation. Information regarding the stimulation parameters used for each participant during void attempts is shown in Table 2. The mean voiding efficiency was 89.2% (SD = 14.5%) with stimulation and 92.1% (SD = 9.1%) without stimulation (p = 0.76). The mean maximum flow rate was 23.3 mL/sec (SD = 12.6) with stimulation and 20.3 mL/sec (SD = 7.28) without stimulation (p = 0.45). Figure 4A shows a void with stimulation and Figure 4B displays the LUT pressure profile before, during, and after the void. A void from the same participant without stimulation is in supplemental Figure 1. Three participants who voided with and without stimulation had a pressure sensor in the bladder. In these cases, at the maximum flow rate the mean bladder pressure change during stimulation was 23.3 cmH2O (SD = 12.6) and without stimulation was 20.3 cmH2O (SD = 7.8), which was not significantly different (p = 0.57).

**Figure 4:**
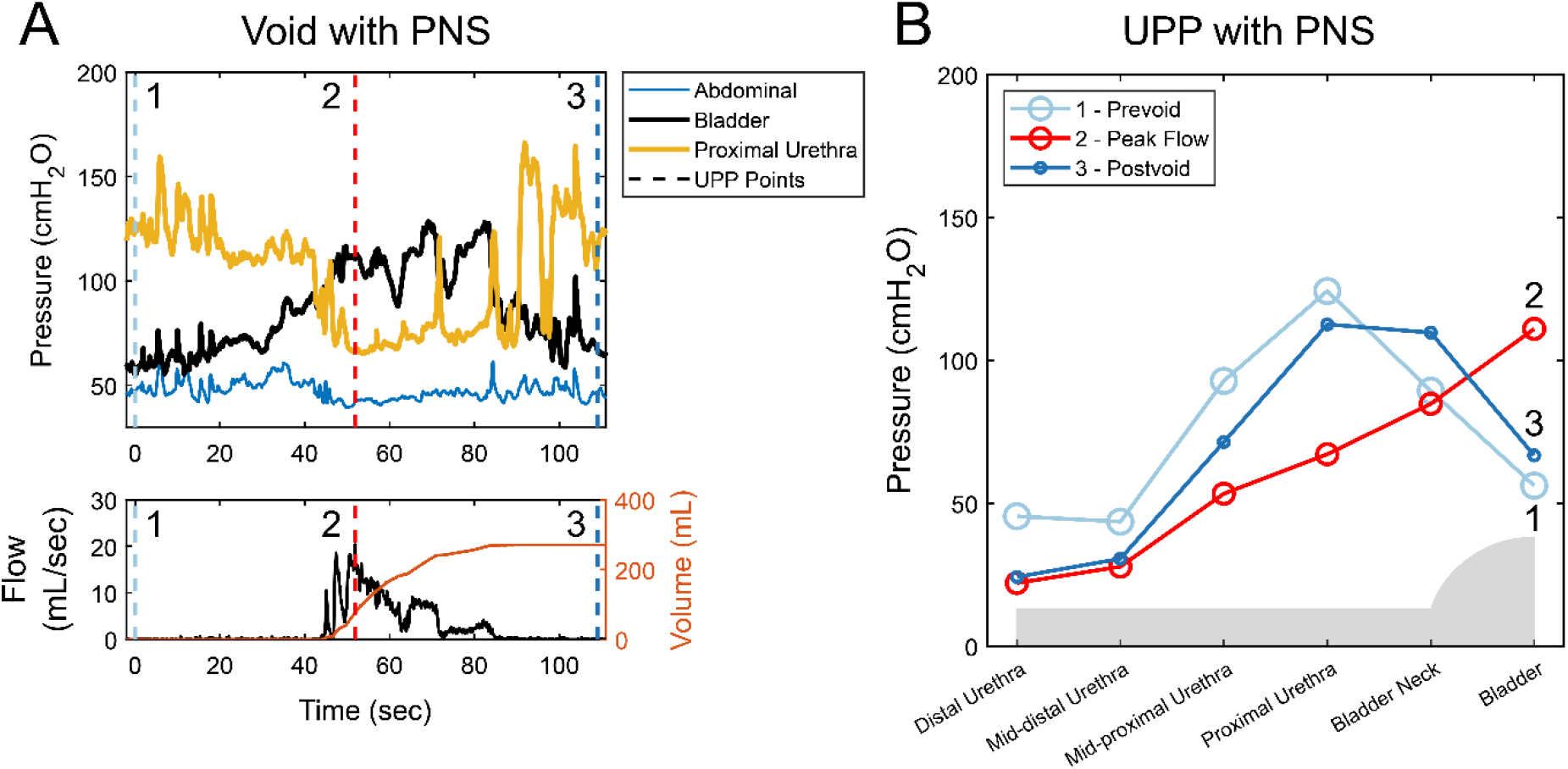
A void with pudendal nerve stimulation (PNS) for participant 1002. A) Pelvic pressures and voiding response during PNS. B) Urethra pressure profiles (UPPs) and bladder pressure from specific time points in A marked with vertical dashed lines.

**Table 2:** Stimulation parameters during void attempts for participants who voided with and without PNS.

| Participant | Electrodes* | Frequency (Hz) | Amplitude | Unit |
| --- | --- | --- | --- | --- |
| 1002 | 0-3+ | 14 | 1.5 | V |
| 1017 | 0-1-3+ | 14 | 0.5 | V |
| 1021 | 0-1-3+ | 14 | 1.5 | V |
| 1024 | 0+2- | 14 | 1.6 | mA |
| 2006 | 2+3- | 14 | NR | V |

\* Electrode definition: Number indicates the active electrode (see Figure 1); +/- indicates anode and cathode(s). Stimulation amplitude was level used by participant at home. NR: not recorded.

## Discussion

Previous studies have shown that PNS can lead to activation of the bladder and urethra in preclinical experiments and in humans under anesthesia or with spinal cord injury (SCI)^6,7,15,17,23–25^. This present study is the first to report urethra and bladder activation from PNS in awake, non-SCI human subjects.

We observed stimulation driven urethra contractions in 11 out of 15 cystometry sessions. Pressure changes occurred along the urethra during high and low frequency stimulation, showing that pudendal stimulation can cause urethra contractions in awake, responsive humans, as previously demonstrated under anesthesia^10^. Urethra contractions were more common with low frequency stimulation than with high frequency stimulation, occurring in more participants and more trials. While low frequency stimulation caused urethra contractions more often than high frequency stimulation, the evoked contractions from high frequency stimulation were generally much larger and tetanic and therefore potentially more useful for preventing urine leakage. High frequency stimulation also had significantly lower MTSAs even though urethra contractions did not affect participant’s tolerance for stimulation. For cases where urethra contractions were not observed it is possible that high frequency stimulation resulted in continuous nerve activation which was typically more noticeable and uncomfortable for participants. This resulted in lower tolerable stimulation amplitudes, and potentially less activation of motor nerve fibers innervating the urethra. Overall, the large number of urethra contractions shows PNS is able to recruit urethra fibers effectively. However as PNS is typically administered before motor fiber activation these results suggest symptom improvement through a mechanism involving activation of sensory afferents similar to sacral neuromodulation^26^.

In one participant, PNS generated excitatory bladder responses in several trials. It is not clear why this participant was the only one with clear PNS-driven bladder responses, but they were especially responsive to stimulation, with a urethra response for 19 out of 21 trials. This might indicate that their implanted lead was particularly successful at recruiting pudendal nerve fibers and was able to influence the lower urinary tract reflexes that influence bladder state^17,27^. It is important to note that in two of the trials with a bladder response the participant noted discomfort with stimulation. The level of stimulation required to activate the bladder likely activates C-fibers as well, potentially explaining why previous reports of bladder pressure changes in response to PNS occurred in people with SCI^15,28^ , preclinical experiments with SCI models^7,19^, or under anesthesia^14,29^. It is possible that other participants had small, stimulation driven increases in bladder pressure during PNS that were indistinguishable from background pressure oscillations. During the stimulation trials, participants were not instructed to void, so descending inhibition of the bladder may have also limited bladder contractions.

We were able to record LUT pressures during bladder voids with and without PNS in a subset of participants. We did not see a significant effect of PNS on voiding function or dynamics. The lack of an immediate change in LUT function suggest that PNS has an indirect effect on LUT function through long term changes in nervous system networks and reflex circuits similar to the proposed mechanisms for sacral neuromodulation^30^. Our findings on the bladder and urethra behavior during voiding matched previous findings^31^. However, using the manometry catheter we observed LUT activity during voids with a level of detail not achieved before. One observation, as visualized in Figure 4B, is that just before voiding starts there is a large decrease in urethra pressure at the location of maximum urethra pressure. This reduction in pressure was fairly localized, only occurring in a region one to two centimeters in length.

Then, during the void, there is a gradual decrease in pressure from the bladder to the end of the distal urethra, showing how LUT pressures aid in urine flow.

Collecting data from awake human participants provided some challenges. Each participant typically sensed PNS. We gradually increased stimulation amplitude as we approached the MTSA. As stimulation increased, baseline pressures could change, or participants could shift positions potentially masking urethra contractions, particularly the sustained contractions associated with high frequency stimulation. Additionally, having two catheters in the urethra made voiding more difficult. While 14 participants attempted to void with both catheters in place, only five participants were able to complete voiding. In some voids the flow urethral pressure profile curve showed signs of restrictive flow, likely from catheterization, potentially masking any effects of stimulation. The low number of completed voids limits the strength of any conclusions on the urethra dynamics during voiding. Future studies could remove the infusion catheter before void attempts to reduce urethra restriction and achieve more consistent voiding.

## Conclusion

This study demonstrates that PNS can evoke urethra and bladder contractions in awake humans which could be useful for future treatments of conditions such as stress urinary incontinence and urinary retention. Additionally, the minimal effect of PNS on bladder or voiding function suggests a PNS mechanism acting through long term changes to bladder neural control circuits. Future studies should investigate whether generated urethra contractions are sufficient to prevent SUI leakage.

## Acknowledgements

Thank you to Vanessa Pruitt and Mackenzie Moore for their efforts in patient recruitment. We are also thankful to the University of Michigan clinical staff for assistance during urodynamic procedures.

## Data availability statement

The dataset used in this study is publicly available on the Pennsieve data sharing portal at: https://doi.org/10.26275/pc8r-r3iu.

## Ethics of approval statement

Approval for this study was obtained from the University of Michigan Institutional Review Board (HUM00165005 & HUM00180124) and was registered on ClinicalTrials.gov (NCT04236596 & NCT04473469) before commencement of study procedures.

## Clinical trial registration

This study was registered at clincaltrials.gov (NCT04236596 & NCT04473469).

