## Supplemental for "Pudendal nerve stimulation recruits the urethra during awake human cystometry"

### Supplemental Information

Supplemental Table 1. Median urethra pressure changes in response to low and high frequency stimulation.

| | Low frequency Trials | | | High frequency Trials | | |
|---|---|---|---|---|---|---|
| Participant | Responsive Trials / Total Trials | Median Distal Pressure Change ($cmH_2O$) | Median Proximal Pressure Change ($cmH_2O$) | Responsive Trials / Total Trials | Median Distal Pressure Change ($cmH_2O$) | Median Proximal Pressure Change ($cmH_2O$) |
| 1002 | NA | NA | NA | 3/8 | 5.0 | 16.3 |
| 1008 | 1/3 | 1.5 | 0 | 1/3 | 10.8 | 9.7 |
| 1010 | 0/5 | 0 | 0 | 0/1 | 0 | 0 |
| 1012 | 0/4 | 0 | 0 | 0/5 | 0 | 0 |
| 1016 | 0/3 | 0 | 0 | 3/4 | 8.1 | 10.9 |
| 1017 | 2/5 | -0.7 | 0 | 0/4 | 0 | 0 |
| 1019 | 0/3 | 0 | 0 | 0/3 | 0 | 0 |
| 1021 | 0/4 | 0 | 0 | 0/5 | 0 | 0 |
| 1022 | 4/4 | 6.0 | 1.5 | 0/4 | 0 | 0 |
| 1024 | 6/6 | 0.4 | 2.0 | 0/6 | 0 | 0 |
| 2001 | 10/11 | 2.5 | 8.3 | 9/10 | 44.9 | 55.6 |
| 2003 | 4/4 | 3.5 | -4.2 | 3/4 | 5.8 | 21.3 |
| 2004 | 1/2 | 3.3 | 4.9 | 2/5 | 34.1 | 63.7 |
| 2005 | 1/4 | 0.9 | 0 | 0/4 | 0 | 0 |
| 2006 | 2/4 | 1.7 | 0.8 | 3/5 | 21.8 | 26.0 |

NA = not applicable, as Participant 1002 had no trials with low frequency stimulation.

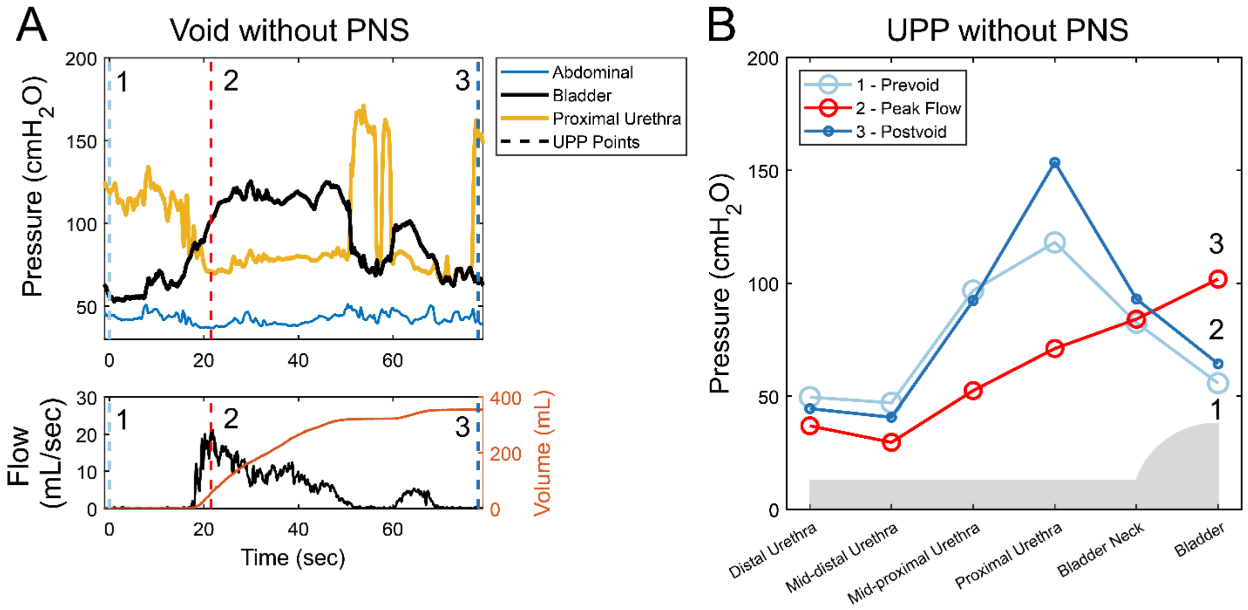


Supplemental Figure 1: A void without pudendal nerve stimulation (PNS) for participant 1002. A) Pelvic pressures and voiding response during PNS. B) Urethra pressure profiles (UPPs) and bladder pressure from specific time points in A marked with vertical dashed lines.
